# Socio-demographic, not environmental, risk factors explain fine-scale spatial patterns of diarrheal disease in Ifanadiana, rural Madagascar

**DOI:** 10.1101/2020.04.02.20051151

**Authors:** Michelle V Evans, Matthew H Bonds, Laura F Cordier, John M Drake, Felana Ihantamalala, Justin Haruna, Ann C Miller, Courtney C Murdock, Marius Randriamanambtsoa, Estelle M Raza-Fanomezanjanahary, Bénédicte R. Razafinjato, Andres Garchitorena

**Affiliations:** Odum School of Ecology, University of Georgia, Athens GA USA; Center for Ecology of Infectious Diseases, University of Georgia, Athens GA USA; Department of Global Health and Social Medicine, Harvard Medical School, Boston MA USA; PIVOT, Ranomafana, Madagascar/ Boston MA USA; Department of Infectious Diseases, College of Veterinary Medicine, University of Georgia, Athens GA USA; Center for Tropical and Emerging Global Diseases, University of Georgia, Athens GA USA; Center for Vaccines and Immunology, University of Georgia, Athens GA USA; River Basin Center, University of Georgia, Athens GA USA; National Institute of Statistics, Madagascar; Ministry of Health, Madagascar; MIVEGEC, Univ. Montpellier, CNRS, IRD, Montpellier, France

**Keywords:** diarrheal disease, precision health mapping

## Abstract

Diarrheal disease (DD) is responsible for over 700,000 child deaths annually, the majority in the tropics. Due to its strong environmental signature, DD is amenable to precision health mapping, a technique that leverages spatial relationships between socio-ecological variables and disease to predict hotspots of disease risk. However, precision health mapping tends to rely heavily on data collected at coarse spatial scales over large spatial extents. There is little evidence that such methods produce operationally-relevant predictions at sufficiently fine enough spatio-temporal scales (e.g. village level) to improve local health outcomes. Here, we use two fine-scale health datasets (<5 km) collected from a health system strengthening initiative in Ifanadiana, Madagascar and identify socio-ecological covariates associated with childhood DD. We constructed generalized linear mixed models including socio-demographic, climatic, and landcover variables and estimated variable importance via multi-model inference. We find that socio-demographic variables, and not environmental variables, are strong predictors of the spatial distribution of disease risk at both an individual and commune-level spatial scale. Specifically, a child’s age, sex, and household wealth were the primary determinants of disease. Climatic variables predicted strong seasonality in DD, with the highest incidence in the colder, drier months of the austral winter, but did not predict spatial patterns in disease. Importantly, our models account for less than half of the total variation in disease incidence, suggesting that the socio-ecological covariates identified as important via global precision health mapping efforts have reduced explanatory power at the local scale. More research is needed to better define the set of conditions under which the application of precision health mapping can be operationally useful to local public health professionals.

## Introduction

Over 700,000 child deaths are attributed to diarrheal disease (DD) annually^1^. The burden of DD is unequally distributed across the globe: 73% of deaths occur in just fifteen low income countries, driven by inequalities in water and sanitation infrastructure and environmental conditions^2^. Understanding risk factors of spatio-temporal patterns of DD is instrumental in designing public health interventions that target populations most at risk. Precision health mapping is an approach that incorporates increasingly available fine-scale social and environmental information into spatial models to explain and predict spatial disease patterns at resolutions finer than those previously possible^3^. This approach has recently emerged as a method to identify areas and populations at risk of disease, and has been successfully used to map the global distribution of diseases with strong environmental signatures, such as mosquito-borne (e.g. malaria^4^, dengue^5^, lymphatic filariasis^6^) and water-borne diseases (e.g. schistosomiasis^7^). There are few examples, however, of efforts that are precise enough and appropriately integrated with implementation to be able to improve local public health strategies.

Diarrheal disease is ideal for precision health mapping because the determinants of environmental suitability for a diarrheal pathogen, such as climate^8,9^, land cover^10^, and soil characteristics^11^, are well known. Hydrological networks, as well as infrastructure and WASH (water, sanitation, and hygiene) practices influence transmission dynamics and individual risk of DD^1012,13^. Upstream land cover has been shown to predict DD prevalence in rural areas of the tropics^10^, with cumulative effects for populations that are downstream of sources of water contamination (e.g. livestock or agricultural run-off^14^). Global analyses of coastal *Vibrio cholera*, the causative agent of cholera, for example, have found that chlorophyll levels, a common consequence of runoff-induced algal blooms, are the strongest predictor of the pathogen’s presence^15^.

Such studies tend to rely on data extracted from large national surveys, such as Demographic and Health Surveys, that are powered to estimate indicators at broad spatial and temporal scales, such as a country or region every five years. When projected to more granular geospatial data, they produce fitted values on the assumption that relationships found at broad spatial scales exist at fine spatial scales. However, because these localized predictions are not typically fitted with localized health data at fine spatio-temporal scales, they may fail to explain patterns of disease at local scales, limiting their ability to inform priorities set by local health actors.

Socio-ecological determinants of DD may be region- and pathogen-specific, so that relationships identified at the global level may not hold locally. For example, despite a proven biological pathway of fecal-oral transmission, the effects of WASH interventions on DD prevalence are ambiguous due to differences in socio-ecological context and specific etiological agents^16^. Pathogens’ temperature responses differ between viral (higher survival in colder environments) and bacterial agents (lower growth in colder environments)^17^, with net effects on DD dynamics dependent on the prevailing set of pathogens. Additionally, precipitation leads to the highest disease prevalence at extremes (i.e. droughts and flooding)^18^, a non-linearity that may be difficult to identify at the local level due to lower variability. At the national and global scale, increased forest cover is consistently associated with lower incidence of DD^10,19,20^, but these associations have not been tested at the fine spatial scale relevant for local action. Addressing this difficulty in downscaling precision health mapping tools is central to evaluating their potential utility.

Here, we investigated the potential for precision health mapping of DD at a fine scale (village-level) in the rural health district of Ifanadiana, in southeastern Madagascar. We leveraged multiple spatio-temporal datasets, including a district-representative longitudinal cohort study and health center case reports in Ifanadiana, to identify the socio-ecological risk factors of DD. We then assessed our ability to predict disease risk at a scale relevant to public health managers. The district comprises a protected tropical rainforest (Ranomafana National Park, a UNESCO World Heritage Site), large areas of agricultural land, and a steep east-west elevation gradient. Due to widespread poverty and low access to improved WASH infrastructure, the population has high exposure to diarrheal pathogens, with children under the age of five and infant mortality rates at twice the national estimates^21,22^. Because of the data availability, the region’s environmental variability, and the population’s high exposure rates, Ifanadiana should be well-suited for the application and validation of precision health mapping at a local scale.

## Methods

### Study area

Ifanadiana district encompasses an area of 3970 km^2^ in the Vatovavy-Fitovinany region of Southeastern Madagascar and consists of about 210,000 people across 13 communes. The majority of the district is rural and the dominant land cover is agricultural land for rice production. The western border includes Ranomafana National Park, a 416 km^2^ protected tropical rainforest. The east-west elevation gradient slopes downward from an altitude of 1400m in the western border to less than 100m in the eastern border. This combination of densely forested area, agricultural fields, and human-developed lands offers an ideal study system to explore how human-modified landcover influences diarrheal pathogen transmission.

Beginning in 2014, the non-governmental organization PIVOT partnered with the Ministry of Health (MoH) to establish a district-level model health system^23^. The intervention strengthens the public health system through a set of programs focused on improving system readiness (infrastructure, personnel, equipment, supply chain), clinical programs (maternal and child health, emergency care, infectious diseases), and integrated information systems at all levels of the district health system (community health, primary care centers, and hospital). Initially started in four communes in 2014, the intervention has since reached six of 13 communes (as of 2019) and will expand to the whole of the district by the end of 2020. In addition, a pilot initiative is underway to upgrade the community health system to support professionalized proactive community case management (Pro-CCM)^24^, in which community health workers visit each household monthly and are able treat DD, malaria, and respiratory infections for children under five to reduce geographic accessibility challenges. Central to this agenda is optimizing the interventions geographically in the context of heterogeneous disease burdens; the work described in this study was done in support of these initiatives.

### Longitudinal cohort dataset

A longitudinal cohort study was conducted to obtain demographic, health, and socioeconomic information from a representative sample of 1600 households in Ifanadiana at two-year intervals (April/May 2014, August/September 2016, and April/May 2018)^21^. The surveys were largely adapted from Demographic and Health Survey (DHS) questionnaires and carried out by the Madagascar National Institute of Statistics (INSTAT). Surveys were stratified across the initial catchment and non-catchment areas of the MoH-PIVOT intervention and divided into 80 clusters, half in each stratum. In each cluster, 20 households were selected at random to be surveyed. The longitudinal study was reviewed and approved by the Madagascar National Ethics Committee and the Harvard Medical School IRB, and de-identified data were provided by the authors for the current study. The current analysis was ruled non-human subjects work by the University of Georgia IRB.

We chose a subset of covariates to include in our models of DD risk, based on proposed drivers and correlates of diarrhea in the literature and exploratory analysis of the full dataset. Only children aged five and under were included in the analysis, and responses were collected from surveys conducted with each child’s mother. A child was determined positive for DD if diarrheal symptoms had been reported in the two weeks prior to the survey date. Demographic data included the child’s sex and age in years. Socioeconomic data included household wealth, mother’s education, defecation practices (e.g. open defecation), and access to an improved water source. The conventional DHS wealth score calculation includes the two WASH variables that are already included individually in our model (access to an improved water source and defecation practices)^25^. To understand the separate influence of these WASH practices, we created a new wealth score for each survey year following standard DHS methods (i.e. principal components analysis) that did not include these variables. This index had high correspondence with the original wealth scores calculated by INSTAT (Pearson’s *r* between 0.941 – 0.989). Wealth scores were transformed into a wealth index by dividing households into five equal quintiles (1^st^ being lowest and 5^th^ being highest) for each survey period. Because wealth indices can serve as a proxy for multiple transmission factors^26^, we conducted a supplementary analysis in which we divided the wealth index into an economic index and a housing index. This separation allowed us to consider the impact of socio-economic status separately from increased exposure to pathogens due to poor housing quality^27^. Finally, exploratory analyses suggested that both age and wealth had non-linear effects on DD risk, leading to the inclusion of quadratic terms in the global model for these two covariates.

### Health system dataset

We collected aggregated monthly health data from the MoH from all public primary care centers (‘*Centre de Santé de Base’*, CSBs) in Ifanadiana. Seven communes have two CSBs and six communes have one CSB, for a total of 20 CSBs across the district of Ifanadiana. The CSBs aggregate the number of all-cause diarrheal cases in children under five years of age on a monthly basis. For communes with two CSBs, the reported cases were combined to result in one monthly incidence value per commune per month. To calculate the incidence of DD per commune, we divided the number of cases by the estimated population of children under five years old. Population values per commune were obtained from official MoH records, which assume a population-wide growth rate of 2.8% per year and a constant age stratification with 18% of the population under the age of five^22^. The study period for this analysis encompasses June 2015 – December 2018, a total of 42 months.

As with all health system datasets, reporting bias is likely to be present because the data only include healthcare-seeking diarrhea cases. While there were differences in healthcare seeking behaviors across communes in the cohort survey dataset, these differences were inconsistent across survey years (i.e. variation in health seeking behavior introduces noise into the data, but not systematic bias) (Figure S2). Only 25.8% of diarrhea cases reported seeking healthcare at a CSB, and so the health system data are likely an underestimate of true disease incidence in the population. As the removal of user fees and other programs implemented in CSBs supported by PIVOT have been shown to increase primary healthcare utilization^22^, we included PIVOT support over time as a binary variable to control for differences in healthcare utilization between CSBs supported and not supported by PIVOT.

### Climate covariates

We used land surface temperature data collected by the MODIS TERRA satellite (MOD11A2) for the date range October 2013 – December 2018, downloaded using the MODIS package^28^ in R v. 3.5.2^29^. This dataset is collected every eight days at a 1 km resolution. Pixels with cloud-cover were removed from the temperature dataset and replaced with spatio-temporally permuted values using the gapfill package^30^ in R v. 3.5.2^29^. The cloud-corrected eight-day data were then aggregated to a monthly estimate by taking the monthly-mean of the eight-day values. We used the Famine Early Warning Systems Network (FEWS-NET) precipitation dataset, created by USAID, to calculate monthly precipitation from October 2013 – December 2018^31^. The precipitation data are available at a 10-day frequency at a 5 × 5km resolution, and were summed by month, resulting in total monthly rainfall in mm. For the cohort dataset, cluster-level climate data were obtained by taking the spatial mean within a 1 km buffer surrounding each village in the cluster. For the CSB dataset, we used the spatial mean across each commune. Temporally-lagged climate variables can represent effects that act over longer time scales and have been shown to be important for DD^18^ and other water-borne diseases^32^. Lags in temperature of up to three months were explored. However, given the high correlation between these values, we chose to include only a one-month lag to reduce collinearity in the model. In the case of precipitation, cumulative values can describe surface water accumulation as a result of seasonal rainfall trends. Therefore, we included one-month lagged rainfall and six-month cumulative rainfall.

### Landcover variables

We created a fine-resolution landcover dataset by processing satellite and OpenStreetMap data. To identify the forest and rice field landcovers, we classified 10 × 10m Sentinel-2 imagery from August 2018 into five land classes (developed, forest, surface water, savanna, and rice fields) using a semi-supervised random forest algorithm, implemented via the Dzetsaka plugin in QGIS^33^ (method described fully in Ihantamalala et al. 2020^34^). This model was trained on 60% of the manually identified regions of interest and validated on the remaining 40% (Kappa = 0.95), and additional points were verified through ground-truthing (62 points) and manual identification via GoogleEarth (254 points). The developed landcover layer was created from residential areas identified in OpenStreetMap data, accessed on April 10 2019 via the Overpass-Turbo API (https://overpass-turbo.eu/). All buildings and residential areas in Ifanadiana district were mapped and verified through a collaborative effort via the Humanitarian OpenStreetMap Team, using satellite imagery and tools available on OpenStreetMap. Polygons were rasterized into a 10 × 10 m resolution to match other landcover datasets.

A large portion of households surveyed in the cohort obtained their water from surface water sources compared to other water sources (46%), and surface water is vulnerable to contamination from upstream pollutants^14^. Therefore, we included the percentage of each type of upstream landcover for streams within 1 km of a surveyed village as a covariate. Forest and rice landcovers were resampled to a 30 × 30 m resolution from a 10 × 10 m resolution by taking the mode of the landcover types within the coarser resolution pixel. Developed landcover was resampled by summation, to give a percentage of developed landcover within each 30 × 30 m pixel. A flow accumulation model and stream dataset were created from the Shuttle Radar Topography Mission Digital Elevation Model^35^ at a 30 × 30 m resolution with a threshold minimum drainage basin size of 1 km^2^. We used this map to calculate the percent of flow accumulation at each stream pixel that traversed each of the three landcover types. All hydrological analyses were done in GRASS 7.6^36^.

### Disease prevalence in the cohort dataset

We used multi-model inference to identify socio-ecological risk factors of an individual child’s risk of DD, or disease prevalence. Multi-model inference allows multiple hypotheses to be considered and incorporates measurements of parameter uncertainty through the process of model averaging^37^. Unlike step-wise approaches that result in one final model, this technique results in a final set of models, as determined by information criteria such as AIC. Model averaging of this final set allows for the calculation of parameter estimates and their uncertainty, as opposed to single valued null hypothesis significance tests. Importantly, this approach is well-suited for the study of socio-ecological systems, often characterized by multiple, interacting variables, due to its ability to identify variables that are consistently strong predictors of the response variable across models^38^. The initial global model was a generalized linear model with hierarchical random effects of the survey cluster nested within the year of survey, to account for the sampling design of the cohort survey study. A total of six individual or household-level socio-demographic variables and nine cluster-level environmental variables were included in the initial global model (Table 1, Figure 1). Only main effects without interactions were included. All variables were inspected for assumptions of normality, and the six land cover variables were *ln*-transformed. Following Gelman^39^, we centered and standardized our predictor variables to a mean of 0 with 0.5 standard deviation to allow for comparison of parameter estimates during model averaging. The response variable was whether an individual child had an episode of DD in the prior two weeks, as reported by the mother (binary). Therefore, we used a binomial distribution with a logit link (logistic regression). We then generated a full model set consisting of all possible subsets of the global model including up to seven covariates, to reduce singularity within the fitted models. The subset of top models consisted of all models with AIC scores within 2 *ΔAIC* of the best fitting model. An average model was obtained from this subset of top models using the zero method of model averaging^40^, implemented in the AICcmodavg package^41^ in R v. 3.5.2. We assessed model performance using the area under the receiver operator curve (AUC) and Tjur’s coefficient of discrimination, *D*, which are performance metrics suitable for models where the response variable is binomial. AUC balances the true positive rate and false positive rate to return a metric ranging from 0 – 1, where a value of 0.5 is equal to a completely random model. Tjur’s *D* is the average difference of the predicted values for positive and negative cases and is an analog of R^2^, ranging from 0 – 1.^42^.

**Table 1.**
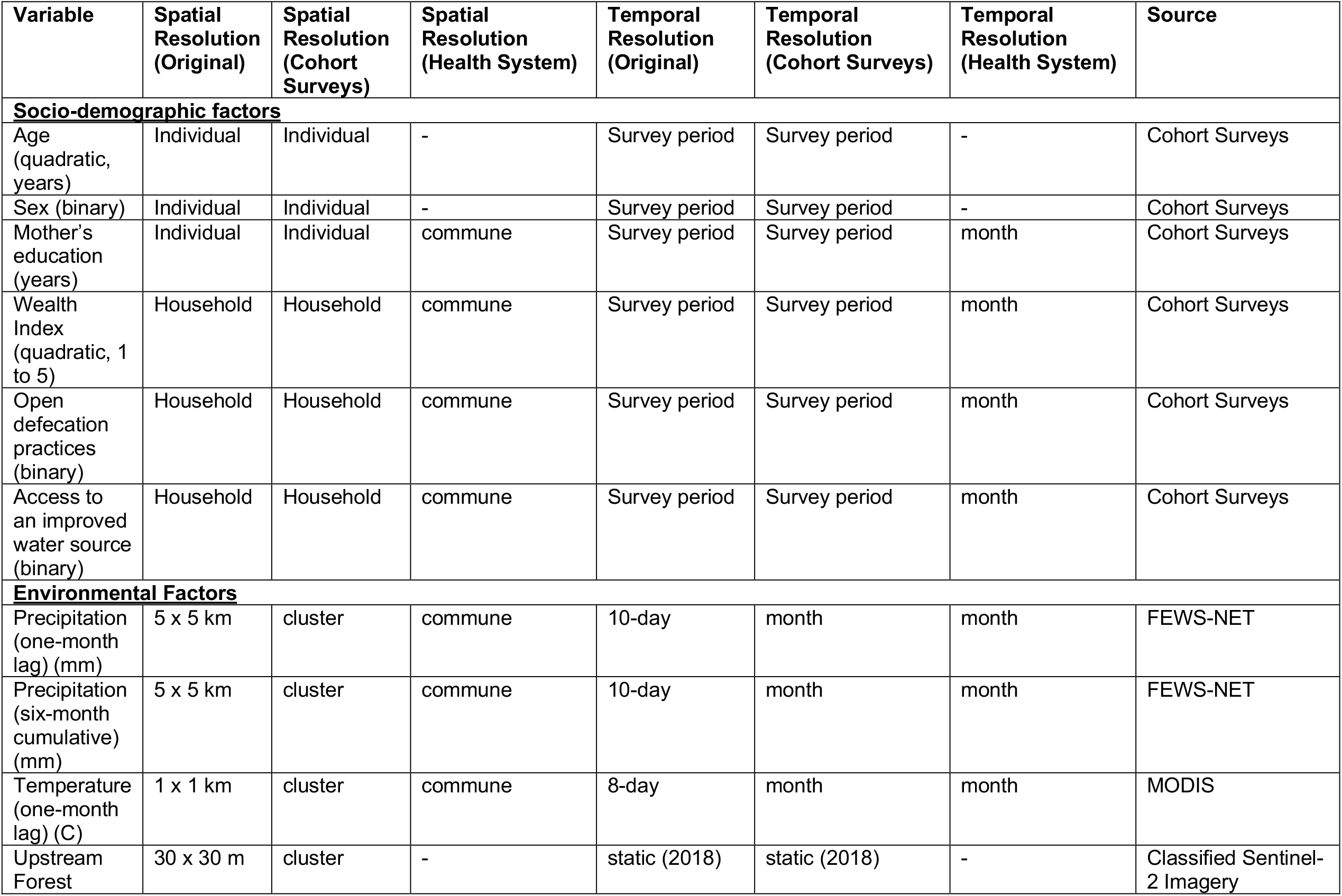
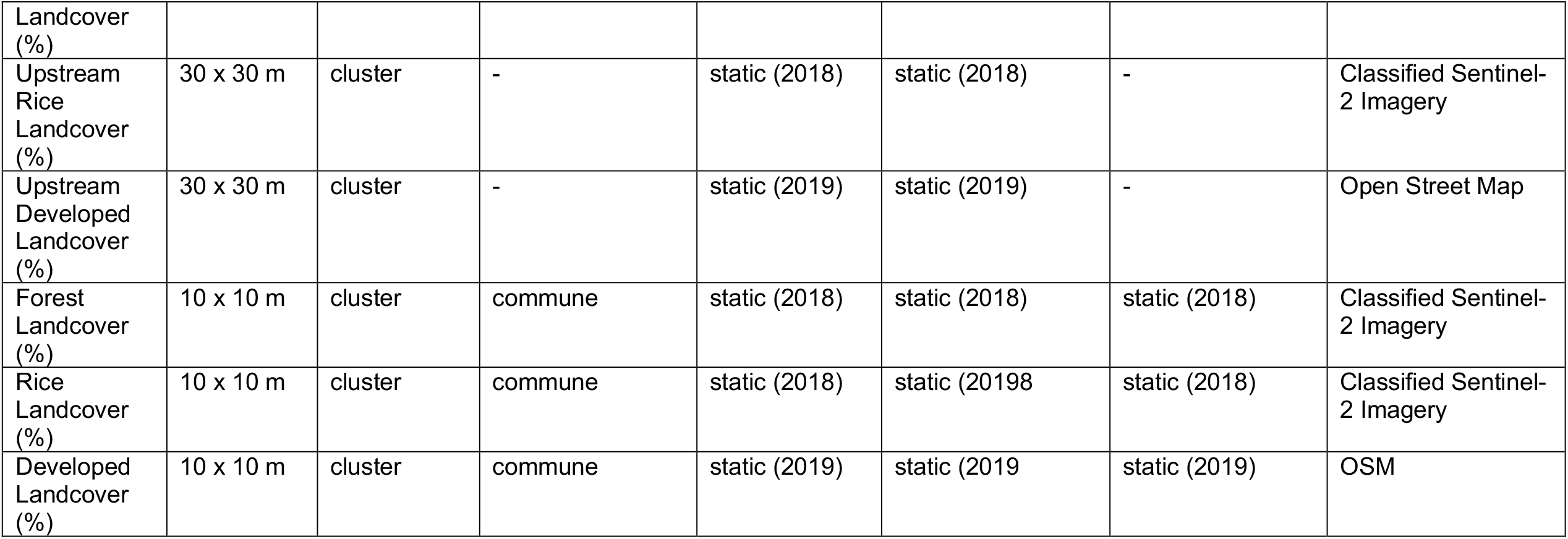
Social and environmental variables used in the models.

**Figure 1.**
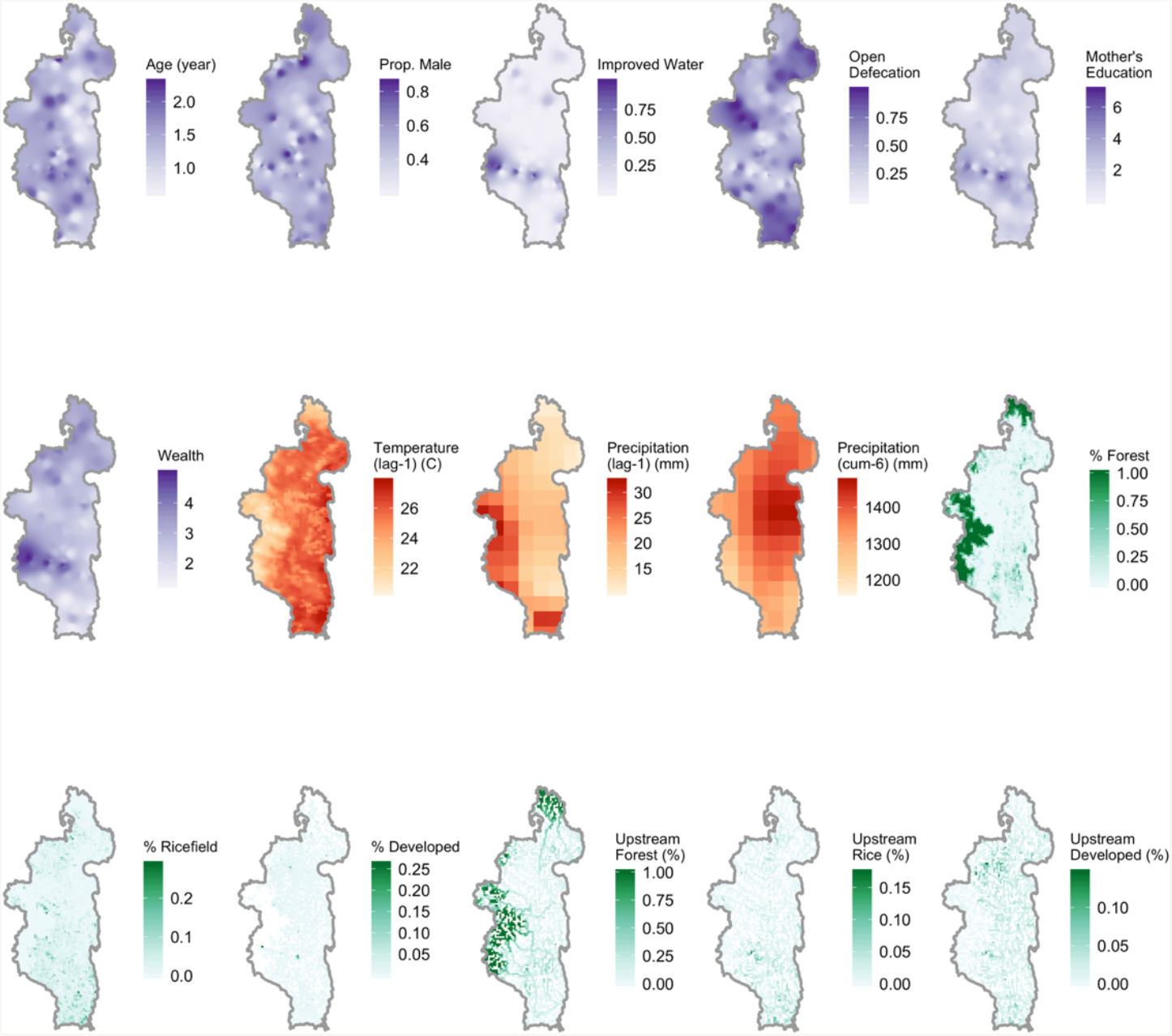
Socio-ecological variables used in the prevalence model mapped across Ifanadiana district. Only data for 2014 is shown. Socio-demographic variables (purple) were interpolated from 80 cluster centroids using third-order inverse distance weighting. Climate variables (red) are shown in their initial spatial resolution. Landcover variables (green) were spatially averaged to 1×1km pixels for visualization.

### Commune-level incidence using health system information

We used a similar multi-model inference approach as described above to explore the socio-ecological risk factors of monthly DD incidence at the level of the commune. The initial global model was fit using a negative binomial distribution and included commune as a random intercept. More complex models including spatially structured random effects and autocorrelation were also considered, but either they offered no improvement on fit (as measured by AIC) over the model with only a random effect of commune or they did not converge (Table S1). There was no evidence of temporal or spatial autocorrelation in model residuals. The model included a subset of the covariates used in the cohort survey analysis. Age and sex were not included because they varied at the level of the individual child and were not spatially structured (Fig. 1). Similarly, the upstream landcover variables were not included because communes are much larger than the watersheds delineated in our flow accumulation model, and so a spatially aggregated summary of such large areas could be misleading. Socio-demographic variables were aggregated to the commune-level by taking the mean of household-level variables from the three cohort surveys and were interpolated linearly to estimate monthly values from June 2015 – December 2018 (Table 1). Discussions with local health workers suggested that DD incidence increased following the national Independence Day (June 26), and so we included the month of July as a covariate in the model. Finally, we included whether or not a commune was part of the PIVOT catchment during that month to control for increased health center utilization following the health system strengthening activities implemented by the MoH and PIVOT in these areas^22^. Because the response variable was incidence per commune, we used different metrics than in the first analysis to assess model performance: root-mean square error (*RMSE*) and Ω^2w^, a form of pseudo-R^2^ suitable for mixed-effects models^43^.

## Results

### Disease prevalence

The cohort survey dataset consisted of 2745 children under the age of five across three sampling periods. There were between four and 30 children in each of the 80 clusters (mean ± *s*.*d*.: 11.45 ± 3.55). District-wide prevalence of DD ranged from 9 – 19% across sampling periods (2014: 145/896, 2016: 127/850, 2018: 84/999), and there was little evidence for spatial correlation in prevalence across clusters (Fig. 2, Moran’s I ranged from 0.00 – 0.06). Additionally, there was low correlation of cluster-level DD prevalence between years (Spearman’s *ρ*: 2014/2016: 0.270, 2014/2018: 0.077, 2016/2018: 0.123).

**Figure 2.**
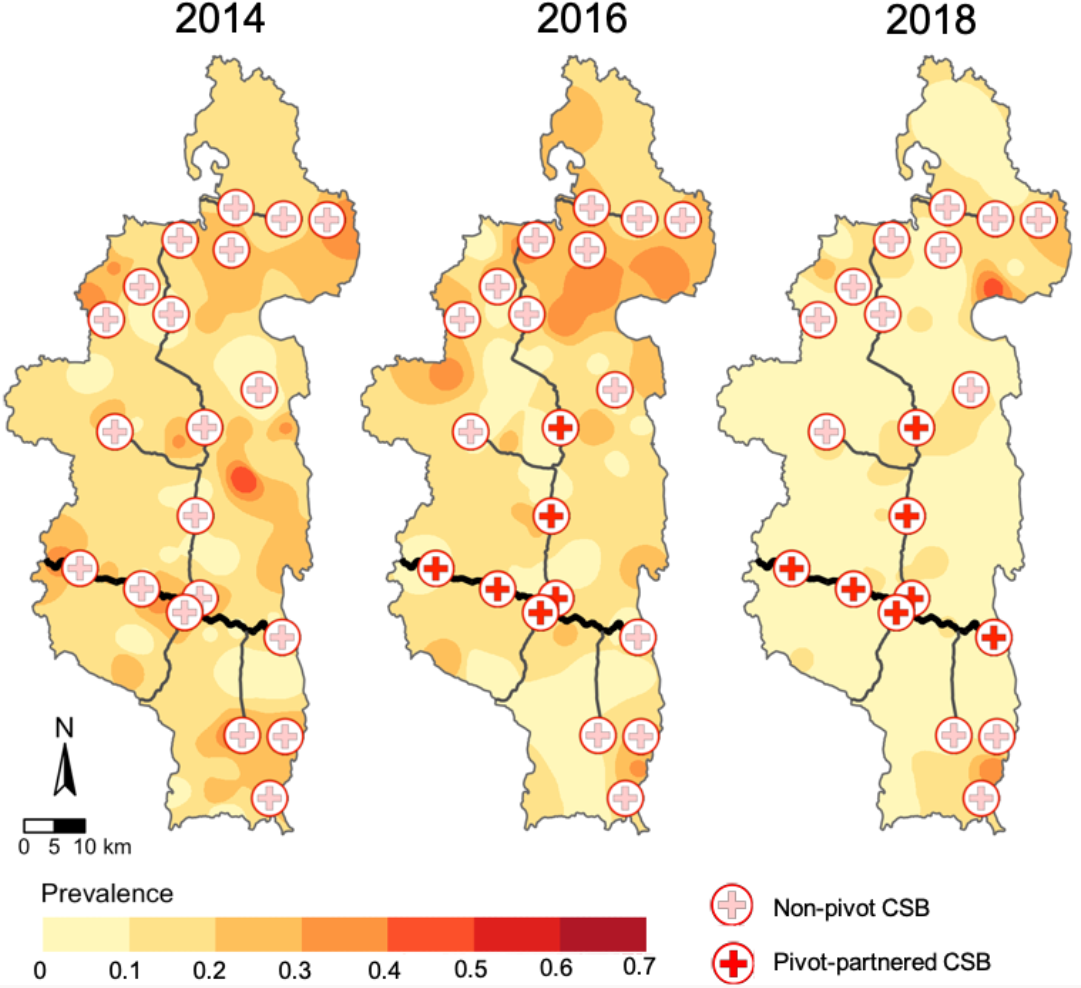
Diarrheal disease prevalence was not spatially structured across clusters. Maps illustrate childhood DD prevalence in prior two weeks interpolated from 80 cluster centroids using third-order inverse distance weighting across the three survey years. Public primary care centers (CSBs) are represented by the cross symbols (filled = PIVOT supported) and the major roads are drawn in black (thick = paved).

The top model set consisted of 15 models out of 41,226 total models that were within 2Δ*AIC* of the best fit model (Table S1). The top model set included 12 of the original 15 predictor variables, and three of those (age, sex, and household wealth) were in all 15 top models (Fig 3). For both age and household wealth, a quadratic relationship better explained individual disease risk than a linear relationship (Fig. 3 B, C). The rate of disease increased from age one to two, and then decreased with increasing age. Similarly, disease prevalence was highest in children from households of intermediate wealth (in the 3^rd^ and 4^th^ wealth quintiles). A supplemental analysis investigating the effects of wealth separately for economic variables (e.g. asset, land, and livestock ownership) and housing quality (e.g. flooring, roofing, and wall materials) found that only economic variables were associated with the individual risk of DD, similar to the relationship with the wealth index in the main model (Figure S1). Male children were more likely to have had diarrhea in the prior two weeks than female children (OR=1.35, 1.08-1.70 95% CI). The regression coefficients of all environmental and climatic variables were included in less than half of the models (Fig. 3), suggesting they were not important risk factors of DD. Finally, the fitted model performed poorly, with an AUC of 0.65 and Tjur’s *D* of 0.027^42^.

**Figure 3.**
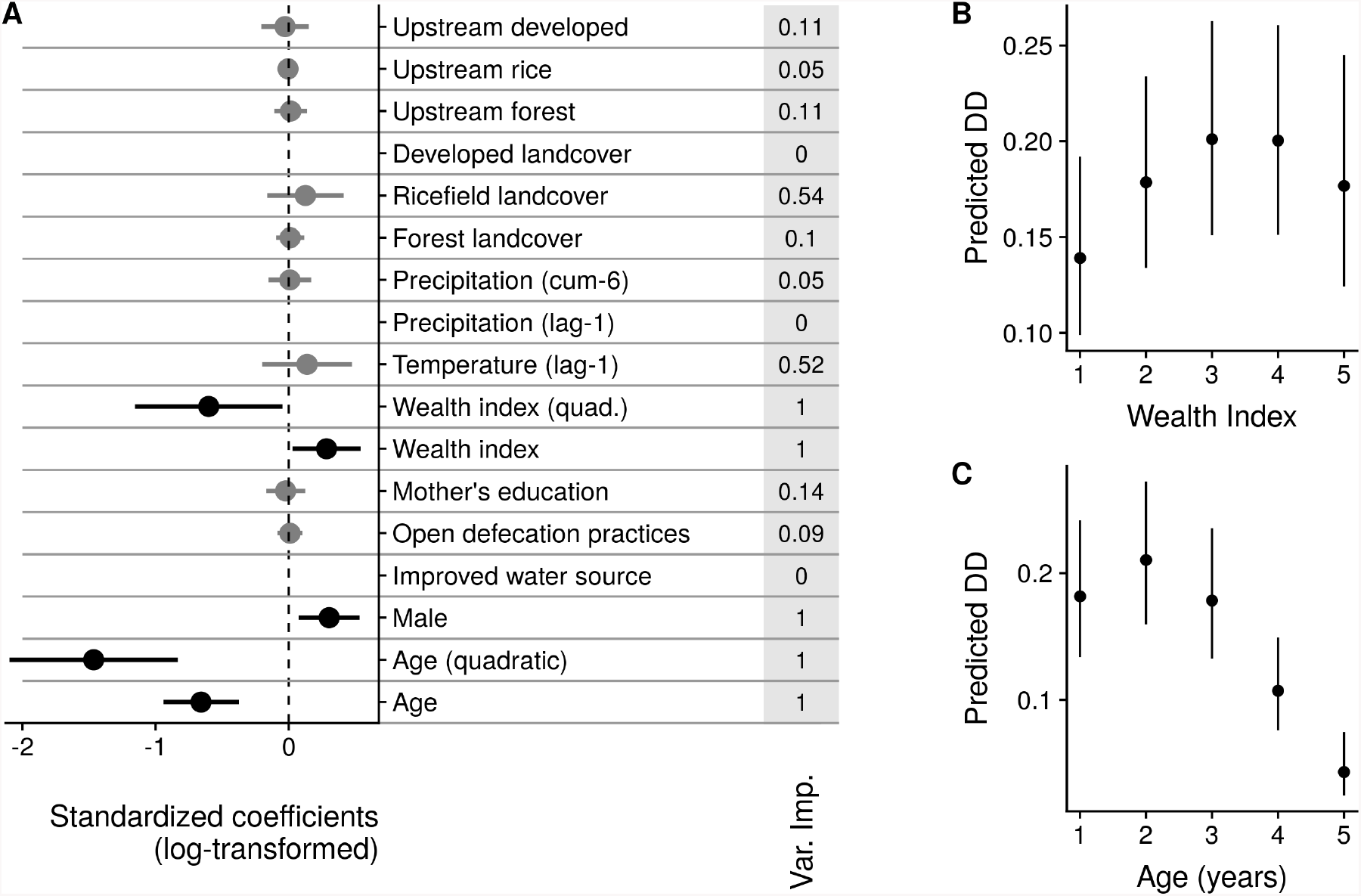
Coefficients from model predicting diarrheal disease prevalence from longitudinal cohort data. A) Log odds-ratios from model predicting DD prevalence from the longitudinal cohort survey data and variable importance scores (in grey column). Variables not included in the final averaged model are not plotted, and variables only included in a subset of the final top model set are plotted in grey. Socio-demographic variables were included in all models and had coefficients with 95%confidence intervals that did not overlap zero. B) Predicted disease risk (mean and 95% CI) for children across wealth indices, holding all other variables constant. C) Predicted disease risk (mean and 95% CI) for children across ages, holding all other variables constant.

### Commune-level incidence

Monthly diarrheal incidence in children under five years of age ranged from 0 to 62 cases per thousand across all 13 communes, with a clear peak in incidence in the winter months (June – August) (Fig. 4). This seasonality was consistent across years. There were differences across communes as well, with Antaretra, Ifanadiana, Tsaratanana, Kelilalina, and Ranomafana, all of which were supported by PIVOT, consistently reporting the highest incidences (Fig. 4). Seven models out of 5,812 were within 2 Δ*AIC* of the top performing model and were included in the averaged model (Table S2). The average model included eleven of the twelve original covariates, with only rice landcover not included in any model. Six covariates (occurrence of the national holiday, a commune falling in the PIVOT catchment, six-month cumulative rainfall, one-month lagged temperature, access to an improved water source, and wealth index) were included in all models and had regression coefficients whose 95% confidence intervals did not include zero. The remaining covariates were only in one model each, and their contribution to the averaged model was very low. The model’s performance was better than that of the cohort analysis model, with a *RMSE* of 5.40 and an Ω^2^ of 0.44.

**Figure 4.**
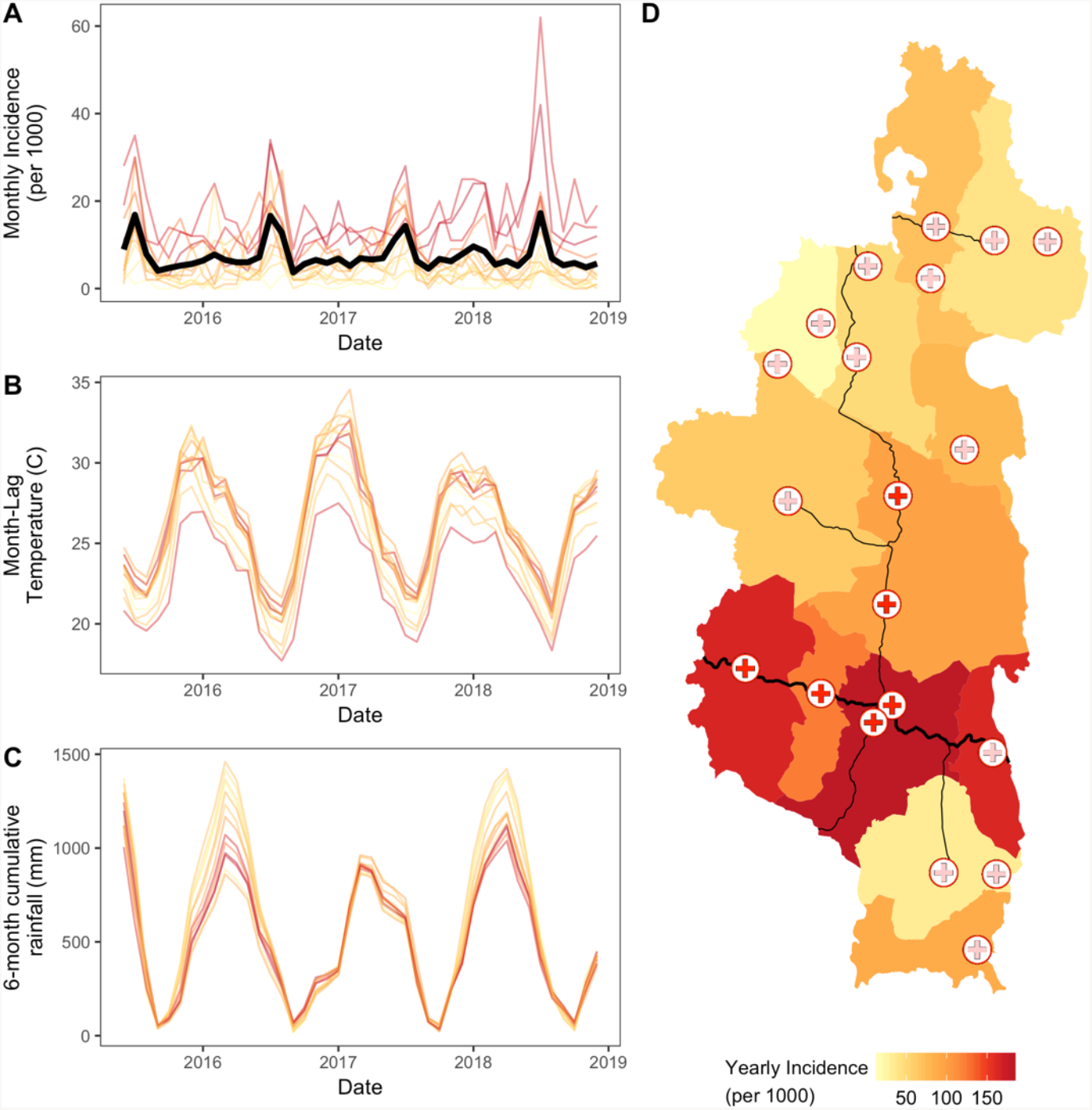
Commune-level diarrheal incidence in children under five across time and space. A) Monthly diarrheal disease incidence for each commune, with the mean incidence across all communes plotted in the bold black line. B) One-month lagged temperature values for each commune. C) Six month cumulative rainfall values for each commune. D) Map of yearly incidence by commune for 2017. Health centers supported by PIVOT are red-filled symbols as in Figure 2. Line colors in A, B, and C correspond to incidence values in D.

As with the analysis of disease prevalence, socio-economic variables had a stronger relationship with disease incidence than environmental variables. DD incidence was lower in wealthier communes (*β* = 0.48, 0.27-0.83 95% CI, Fig. 5). Unlike the prevalence analysis, the quadratic term was not found in the top model set, signifying a linear relationship. Incidence was higher in communes where a larger proportion of the population had access to an improved water source (*β* = 1.85, 1.09 – 3.11 95% CI). One-month lagged temperature and six-month cumulative precipitation influenced seasonal disease incidence, with lower temperatures and higher cumulative rainfall associated with higher incidence rates (Fig. 5; temperature *β* = 0.80, 0.71 – 0.89 95% CI; rainfall *β* = 1.19, 1.08 – 1.32 95% CI). Despite these effects, the standardized regression coefficients were smaller than those of the socio-economic variables. All other environmental variables had low variable importance scores (Fig. 5), suggesting they had little influence on disease incidence.

**Figure 5:**
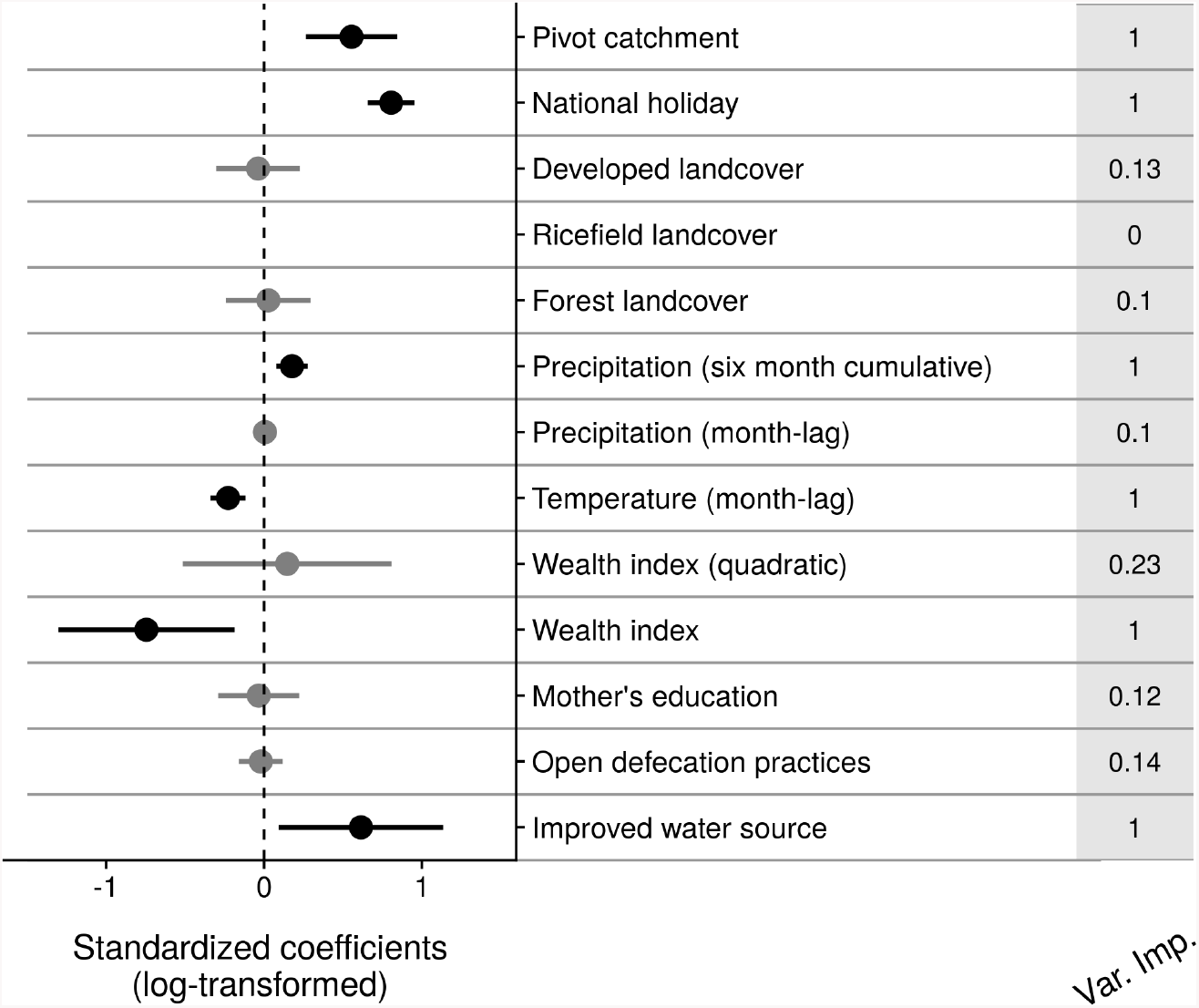
Coefficients from model predicting commune-level diarrheal disease incidence from health system data and variable importance scores (in grey column). Variables not included in the final averaged model are not plotted, and variables only included in a subset of the final top model set are plotted in grey.

## Discussion

There is growing interest in the potential for ‘big data’ to be used to support increasingly optimized and proactive public health interventions. Predicting DD risk through precision health mapping accordingly offers great potential to prevent diarrhea-caused morbidity and mortality. However, there is a substantial gap between our collective analytic capabilities and the use of those capabilities to solve problems where they matter the most – in areas of extreme poverty with high burdens of disease. For precision health mapping to lead to actionable interventions, the global relationships between DD and its socio-ecological risk factors must remain at the fine spatial resolutions relevant to public health actors. Here, we used a multi-dataset analysis to identify social and ecological risk factors of diarrhea in children under five years of age in a rural health district in Madagascar. Environmental variables contributed little to spatial variation in disease incidence and prevalence but did explain the seasonality of diarrheal disease. Socio-demographic variables, however, were predictive of diarrheal disease risk at both the individual and commune levels. This questions whether precision health mapping approaches that rely on environmental and climatic variables can accurately predict disease risk at local scales.

Our results suggest that children of households from the longitudinal study with intermediate wealth scores have a higher risk of diarrhea than those from households with the highest or lowest wealth scores; yet, wealthier communes have lower disease incidence rates. Our cohort findings are contrary to the general consensus that diarrheal prevalence and morbidity is lower in households from upper wealth quintiles^44–46^. However, these studies are often done at a national or multi-national scale, where wealth variability may be larger, and they include both urban and rural populations. When focusing only on a rural population, Kumi-Kyereme and Amo-Adjei^47^ found that the effect of wealth is less clear and can have a positive relationship with diarrheal risk. Given that over 70% of the population in Ifanadiana lives in extreme poverty^21^, it is possible that living conditions of households with intermediate and lower wealth scores were comparable, resulting in similar exposure to diarrheal pathogens. Our analysis of commune-level health center data finds that the population of communes with greater access to an improved water source had higher rates of DD. However, overall access to an improved water source remained relatively low (the highest being 53% of a commune’s population), and most communes had less than 25% of the population using an improved water source. When disease prevalence is high and WASH infrastructure coverage low, improvements to WASH may counterintuitively increase DD risk by concentrating exposure at sanitation points and increasing transmission due to human contact at these points, rather than water-borne transmission routes^48^.

We found evidence for very strong seasonality in DD incidence, which is in agreement with other studies focused on enteric diseases in the tropics^49,50^. DD incidence was higher during the cooler, drier winter months, and our analysis found cooler temperatures the month prior to be associated with increased incidence. Enteric viruses have higher survival and environmental persistence at colder temperatures^11^, which may suggest that enteric viruses are the primary causative agents of DD in Ifanadiana. This is further supported by a study conducted in human populations near Ranomafana National Park that found viral infection rates of over 50% during the austral winter^51^. Colder temperatures during winter months may also be associated with human behavior changes that increase transmission rates, such as indoor crowding. Interestingly, only six-month cumulative rainfall trends influenced disease incidence, and not one-month lagged rainfall. While there is evidence that extreme rainfall events, such as flushing or flooding increase DD in other regions^18,52^, long-term rainfall patterns have a closer relationship with DD than extreme events in Ifanadiana. Virus persistence and infectiousness is higher in higher-moisture soils^17^, and a wetter rainy season could increase exposure to enteric viruses by increasing soil moisture and thereby viral persistence.

We conclude that climatic factors do not contribute significantly to spatial variation in DD risk across Ifanadiana district. Although the district spans an elevational gradient of 100 – 1400 m, its area is only approximately 45 × 125 km at its widest, which may not offer enough climate variation across space to observe a clear spatial trend. For precipitation especially, which is available at a 5 km resolution, there is little variation across clusters within a sampling period (Fig. 1). The collection of finer-scale rainfall or watershed-level discharge data could aid in exploring these relationships further and would reduce known biases of remotely-sensed rainfall data^53^, but unfortunately is not available for this geographic region. At the scale of the administrative district, therefore, the effect of climate is important in contributing to seasonal, but not spatial, trends in DD.

After controlling for seasonal climate variables, the occurrence of the national holiday remains associated with higher DD incidence. Studies on short-term changes in disease dynamics due to seasonal cultural events in low income countries are rare, but changes in contact rates associated with holidays and mass migration have been shown to influence disease transmission in other systems^54,55^. In Ifanadiana, the movement of people associated with the holiday could increase contact rates and increase pressure on existing sanitation systems, leading to increased transmission of DD. Additionally, changes in diet and increased food sharing associated with celebration of the holiday could increase children’s exposure to DD pathogens^56^. The incorporation of cultural event calendars and population movements have led to increased predictive accuracy of the timing of outbreaks of measles in Niger^57^ and Chagas disease in Peru^58^, and their contribution to DD should be explored further in this and other geographical contexts.

Unlike past studies^10,14^, we find little evidence for a spatial pattern in DD associated with nearby or upstream land use. It has been hypothesized that populations downstream of undisturbed forest would have lower risk of DD than those downstream from agricultural or developed areas due to lower contamination of surface water and through the filtering of contaminants^10^. However, the largest forest in Ifanadiana, Ranomafana National Park, attracts thousands of tourists annually. In this case, the additional water contamination due to tourism may negate any beneficial “filtering” mechanisms of the forest. The amount of developed area, a fine-scale proxy for population density, also did not impact disease risk. Ifanadiana is a primarily rural district, with only a few large towns (population greater than 3000 people) located along paved roads. While communes with large towns along the main road did have higher incidence of DD in the health system dataset, this variation was primarily explained by the presence of PIVOT-supported infrastructure and the random effect of commune in the model, not population density (represented by the amount of developed area). Therefore, it is likely that unmeasured characteristics of these communes, such as mobility or water quality, are contributing to their high rates of DD incidence.

In conclusion, we fail to find evidence that precision health mapping accurately describes local patterns in DD risk in this context. Therefore, precision health mapping may not actually be useful for informing local health authorities of target areas for the implementation of public health interventions. Despite the availability of extensive longitudinal epidemiological data and high-resolution landcover and environmental information, we found that our models poorly predicted disease prevalence. Precision health mapping at this scale may require even finer scale spatial data, such as that collected via community health worker programs that combine proactive care with mobile-health data collection. The strongest predictors of disease risk in our context are socio-demographic or cultural (e.g. holidays), but their lack of spatial structure compared to environmental variables may limit the performance of precision health mapping exercises. Precision health mapping should be developed and adapted for different environmental and social contexts in order to better define the set of conditions under which its application at fine-spatial scales can be of use to public health professionals.

## Data Availability

Code to reproduce this analysis will be posted onto figshare upon publication of this manuscript. Environmental covariate data will also be shared openly, but we are not permitted to openly share health system data or data from the longitudinal cohort study.

## Acknowledgements

MVE was supported by an NSF Graduate Research Fellowship while conducting this research. We would like to thank the PIVOT research interns, Mauricianot Randriamijaha, Tanjona Andreambeloson, and Miadana Rakotozafinirainy, for their contribution to the OpenStreetMap data and support on this project. We also thank the PIVOT community team and CSB staff for their help in the collection of the health data.

